# Proximal and Distal Bimanual Motor Skill Learning After Stroke: Impairments, Trajectories, and Clinical Contributors

**DOI:** 10.64898/2026.09.02.26362113

**Authors:** Coralie van Ravestyn, Benoit Bihin, Julien Lambert, Benoit Herman, Frédéric Crevecoeur, Thierry Lejeune, Yves Vandermeeren

**Affiliations:** UCLouvain/CHU UCL Namur (Godinne), Neurology Department, Stroke Unit/Motor Learning Lab, Yvoir, Belgium; UCLouvain, Institute of NeuroScience (IoNS), NEUR Division, Brussels, Belgium; UCLouvain, CHU UCL Namur (Godinne), Scientific Support Unit (USS), Yvoir, Belgium; Institute of Mechanics, Materials and Civil Engineering (iMMC), UCLouvain, Louvain-La-Neuve, Belgium; UCLouvain, Louvain Bionics, Louvain-la-Neuve, Belgium; Institute of Information and Communication Technologies, Electronics and Applied Mathematics (ICTEAM), Université catholique de Louvain, Louvain-la-Neuve, Belgium; UCLouvain, Institut de recherche expérimentale et clinique (IREC), Louvain-la-Neuve, Belgium; Cliniques Universitaires Saint-Luc, Physical Medicine and Rehabilitation Department, Brussels, Belgium

## Abstract

**Background:** Although coordination between the upper limbs (ULs) is essential for daily life activities and may benefit from early post-stroke plasticity, how patients early after stroke achieve bimanual motor skill learning (bim-MSkL) remains poorly understood. We investigated whether acute and subacute stroke patients retained bim-MSkL capacity, whether bimanual control policy and motor sequence were dissociable components, whether bim-MSkL was more impaired in the distal than proximal UL segment, and whether clinical factors predicted bim-MSkL.

**Methods:** Ninety-seven (sub)acute stroke patients and 60 age-matched healthy individuals (HI) trained over three days on proximal and distal bimanual tasks in this randomized trial. Baseline motor and neuropsychological impairments were assessed. On day 3, participants were randomized into two subgroups on the proximal task to dissociate control policy from motor sequence. Learning trajectories, perturbation-related performance disruption, and predictors of improvement were analyzed.

**Results:** Accuracy and coordination improved in patients on both the distal and proximal tasks over three days, although less than in HI, while proximal speed improvement remained limited. Control policy swap induced larger performance disruption than motor sequence change, whereas the relearning slopes did not differ in patients. Unlike HI, patients showed no proximal-to-distal gradient in bim-MSkL despite greater distal than proximal baseline impairment. Baseline tasks performances, subacute motor and attentional impairments were the strongest predictors of bim-MSkL impairment early after stroke.

**Conclusions:** Most patients with mild-to-moderate impairment exhibited substantial residual proximal and distal bimanual motor learning capacity early after stroke. Impairment profiles support individualized bimanual rehabilitation strategies integrating both motor and cognitive demands.

## Introduction

Motor skill learning (MSkL), fundamental for activities of daily living (ADLs)^1,2^, is characterized by rapid initial improvements followed by slower performance gains, long-term retention, generalization of the learned skill to novel contexts, and automaticity^2,3^. Following stroke, MSkL contributes to both spontaneous and rehabilitation-induced recovery by supporting the (re-)acquisition of motor skills^4,5^. However, the mechanisms underlying post-stroke MSkL remain incompletely understood^5^.

Because most ADLs require coordinated use of both upper limbs (ULs), post-stroke recovery depends largely on relearning skilled bimanual behaviors^6^. Although preserved bimanual MSkL (bim-MSkL) has been reported in patients with chronic stroke^7–9^, little is known about bim-MSkL early after stroke, a critical period of heightened neuroplasticity^10, 11^. Moreover, because MSkL depends on both motor execution and cognitive processes, impairments in either domain may limit bim-MSkL early after stroke^12, 13^.

Whether stroke differentially affects proximal (shoulder–arm) and distal (hand–fingers) bim-MSkL also remains unclear. These UL segments rely partly on distinct neural substrates^14^. Although post-stroke recovery is often assumed to progress from proximal to distal segments, supporting evidence remains inconsistent^15, 16^. Determining whether proximal and distal bim-MSkL are differentially affected could improve our understanding of motor recovery and inform targeted rehabilitation strategies^17^.

Robotic technologies enable objective quantification of UL motor performance, interlimb coordination, and motor learning ^18^, while allowing complementary components of bim-MSkL to be examined separately^19^. For example, bim-MSkL requires the acquisition of both a control policy (the sensorimotor mapping between UL movements and task goals) and a motor sequence (the temporal organization of repeated movements)^20^.

Using robotic devices, we investigated proximal and distal bim-MSkL early after stroke. We hypothesized that: (1) proximal and distal bim-MSkL would be impaired compared with healthy individuals (HI); (2) control policy and motor sequence would represent dissociable components of bim-MSkL; (3) distal bim-MSkL would be more severely impaired than proximal bim-MSkL; and (4) motor and cognitive impairments would predict bim-MSkL performance.

## Methods

### Population

The study was approved by the local Ethics Committee and conducted in accordance with the Declaration of Helsinki. The manuscript was prepared in accordance with the STROBE reporting guidelines. The study was registered at ClinicalTrials.gov (NCT05760846).

Eligible participants were HI or patients with a first-ever, unique acute/subacute stroke, aged 40-90 years. For patients, stroke onset had to have occurred between 24 hours and 21 days before enrolment. Exclusion criteria included substance abuse, inability to perform experimental tasks, major cognitive impairment (inability to understand simple commands or severe cognitive deficits, defined as a Montreal Cognitive Assessment (MoCA score <10/30), uncontrolled medical conditions, contraindications to MRI, and other neurological disorders.

To capture a broad spectrum of neurological deficits and bim-MSkL impairments, patients with a first-ever ischemic or hemorrhagic stroke meeting the eligibility criteria were included. Data were managed using REDCap. Sample size was determined a priori based on effect sizes (Cohen’s d) estimated from pilot data, assuming α=0.05 and 80% power, yielding a target sample of 60 patients (30 in each randomization group) and 50 HI.

### Study design

Patients underwent motor, cognitive, and neuropsychological assessments as well as structural MRI during hospitalization at CHU UCL Namur (Godinne, Belgium). HI completed the same experimental protocol except for MRI and most clinical assessments (Fig. 1). Participants trained over three consecutive days (D1-D3) on two bimanual tasks targeting proximal and distal upper-limb function, respectively. For each task, participants completed 10 training blocks per day (T1-T30). On D3, an additional 10 blocks (T31-T40) were performed to assess generalization of the learned skill.

**Figure 1.**
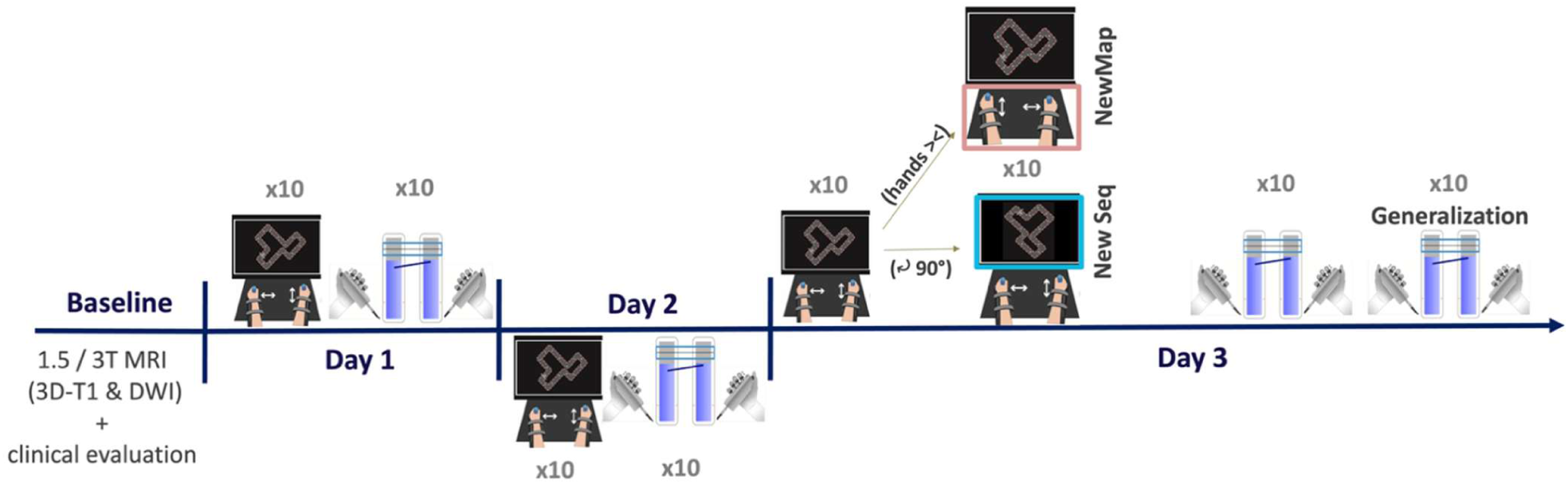
Study design. Participants were trained over three consecutive days on two bimanual tasks: CIRCUIT on the REA²Plan® (proximal bim-MSkL) and LIFT THE TRAY on the Dextrain Manipulandum® (distal bim-MSkL). Ten 1-minute training blocks per day were performed on each task. On D3, participants were randomized into two subgroups to isolate the proximal control policy and motor sequence components by adding ten additional trainings on the CIRCUIT task, with either arms coordination policy reversed (NewMap group, top) or CIRCUIT rotated by 90° while keeping the same arms coordination policy (NewSeq group, bottom). After the 10 training blocks on the LIFT THE TRAY task on D3, 10 additional trainings were assessed with a change in the sequence of force levels to assess generalization. At baseline, patients underwent functional, motor and cognitive assessment scales and MRI. HI followed the same protocol except for MRI and most clinical scales.

### Motor, functional and cognitive assessment

Stroke severity was assessed using the National Institute of Health Stroke Scale (NIHSS)^21^. UL motor and somatosensory impairments were evaluated using the Fugl-Meyer Assessment Upper Extremity (FMA-UE)^22^, whereas functional impairment was assessed using the Shoulder Abduction Finger Extension (SAFE)^23^, Arm Motor Ability Test (AMAT)^24^ and digital dexterity using the Purdue Pegboard Test (PPT)^25^. Global cognitive impairment was evaluated using the MoCA^26^. Attention and visuospatial processing were evaluated using the Bells test^27^ and the Wechsler Adult Intelligence Scale III-Key subtask (Key-WAIS III)^28^. Fatigue was assessed using the Visual Analogue Scale for fatigue (VAS-F)^29^, Stroke lesion volume was measured based on 3D-T1 MRI. HI were only evaluated using the Bells test and Key-WAIS III.

### Experimental devices and tasks

#### CIRCUIT task

On D1, participants underwent active range-of-motion (ROM) calibration to account for inter-individual differences in UL morphology and stroke-related impairment. The robotic workspace was scaled to 80% of each participant’s active UL ROM.

The CIRCUIT task, described previously^30,31^, was used to assess proximal bim-MSkL using the REA²plan® (AXINESIS, Belgium) (Fig. 1). Participants were instructed to complete as many laps as possible during each 1-min training block while keeping the cursor within the track, requiring a continuous trade-off between speed and accuracy. Cursor movement resulted from coordinated UL movements: one arm controlled lateral displacement (left–right axis) and the other controlled anteroposterior displacement (sagittal axis in the horizontal plane)^20,30,31^. Each training block was followed by a 30-s rest period. During the generalization phase on D3, participants were randomized to one of two experimental conditions for 10 additional blocks on D3: (i) New Sequence (NewSeq), in which the circuit was rotated by 90° while preserving the original arms-to-cursor mapping, thereby altering the spatial motor sequence; or (ii) New Mapping (NewMap), in which the arms-to-cursor mapping was reversed, disrupting both the previously learned control policy and the motor sequence.

A second randomization counterbalanced the two possible arm-to-cursor mappings (i.e., which UL controlled each cursor axis) to control for arm dominance, biomechanical asymmetries, directional movement biases, and stroke laterality.

For patients, randomization was stratified RStudio (R Foundation for Statistical Computing, Vienna, Austria; version 4.4.1) according to the FMA-UE score (0–22 / 23–44 / 45–66), time since stroke (week 1 / 2 / 3), stroke laterality (dominant / non-dominant hemisphere) and presence of hemineglect (yes / no). For HI, randomization was stratified according to sex (male / female), age (40–50 / 51–70 / 71–90 years) and education level (<12 years / ≥12 years) were used.

#### LIFT THE TRAY task

Distal bim-MSkL was assessed using a bimanual version of the Dextrain Manipulandum® (Dextrain, France)^32–34^. The system comprised two manipulanda connected to a computer display, each equipped with five force sensors, allowing simultaneous measurement of forces produced by individual fingers. Force data were sampled at 25 Hz and processed offline using custom RStudio scripts.

Participants were instructed to move a virtual tray into a horizontal target zone as quickly as possible and maintain it centered and level within the target by coordinating pinch forces generated by both hands. The vertical position of each tray extremity was controlled by the sum of thumb-index pinch forces generated by the corresponding hand. Consequently, maintaining a horizontal tray required equivalent force production by both hands. Each target force level was maintained for 4 seconds before automatically switching to the next level (Figure S1).

A trial consisted of seven force levels presented in a predefined sequence. The sequence was repeated ten times, separated by 10-second rest periods, to form one training block. After block 30 on D3, participants completed ten additional blocks with a modified force sequence to assess motor sequence generalization.

### Data Analysis

#### CIRCUIT task

Proximal bim-MSkL was quantified using Speed and Accuracy and a Bimanual Coordination Factor (BCF) as as primary outcomes. All metrics were computed over 3-s epochs, resulting in 20 bins per 1-minute training block. Speed (cm/s) was defined as the common cursor velocity and Error (1/cm) as the area between the actual cursor’s trajectory and the ideal trajectory (i.e., the circuit midline). To reduce the influence of outliers, Speed and Error were log-transformed. Error was subsequently sign-reversed to derive Accuracy (-Error, arbitrary units (a.u.)). BCF [a.u.] quantified bimanual coordination by assessing the similarity and synchrony of movement velocities between the two ULs. Specifically, BCF measures phase coherence between upper-limb velocities and was calculated as follows^30,31^:

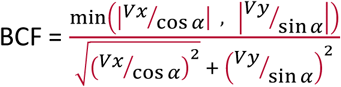

where *V*_x_ and *V*_y_ denote upper-limb velocities along the X-and Y-axes, respectively, and α represents the angle between the start and end points relative to the horizontal plane. The numerator corresponds to the minimum velocity component, whereas the denominator represents total bimanual velocity. Perfect synchronization of both UL yields a theoretical upper bound of 1^31^.

Speed, Accuracy, and BCF were averaged across each 1-minute training block.

#### LIFT THE TRAY task

Distal bim-MSkL was quantified using Dext-Accuracy and the Bimanual Coordination Coefficient (BICO) as the primary outcomes. Pinch force was calculated separately for each hand as the sum of thumb and index finger forces (N). For each side, error was computed as the difference between target force and applied force.

When left-and right-sided errors shared the same sign, the total error corresponded to their mean, representing a uniform translational deviation of the tray from the target position (Figure S1A):

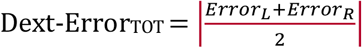

When left-and right-sided errors had opposite signs, indicating tray tilt, total error was computed from the geometric deviation of the tray relative to the target position (Fig. S1B):

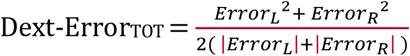

This approach accounts for rotational deviation while avoiding overestimation of global trajectory error when opposing force deviations partially compensate for one another.

BICO was calculated as the difference between the absolute force magnitudes generated by the left and right hands (N):

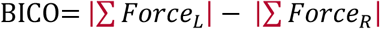

The raw BICO value represents force asymmetry between the two hands. For longitudinal analyses, the sign was transformed such that increasing values consistently reflected improved bimanual symmetry and coordination, thereby matching the directional convention of the other performance measures.

Dext-Error and BICO were computed at the sampling frequency (25 Hz) and averaged across the 4-second duration of each target force level. Both variables were log-transformed to reduce the influence of outliers. Dext-Error was subsequently sign-reversed to derive Dext-Accuracy (−Dext-Error; a.u.), which was used for statistical analyses.

For both tasks, baseline performance was defined as the first training block (D1T1). Overnight retention was quantified as the change between T10 and T11 (D1-D2) and between T20 and T21 (D2-D3). Total learning was defined as the performance change between T1 and T30. Following introduction of the task perturbation on D3, performance disruption (DROP) was defined as the difference between T30 and T31, whereas relearning/generalization was quantified over T31-T40.

### Statistical analyses

All analyses were performed in RStudio (R Foundation for Statistical Computing, Vienna, Austria); analysis code is available upon request.

Proximal and distal outcomes were normalized as z-scores relative to baseline performance in HI (D1T1).

Between-group differences at baseline (T1) and end of training (T30) were assessed using linear models. Learning trajectories were analyzed using linear mixed-effects models (LMM) including training block, group, and their interaction as fixed effects, with participant-specific intercepts and slopes for training block to account for repeated measures and inter-individual variability. Differences in bim-MSkL improvement between groups were assessed from the group × training block interaction term. β denotes estimated regression coefficients derived from linear models or LMM. Because outcome measures represented distinct, predefined dimensions of MSkL and were analyzed independently, no global correction for multiple comparisons was applied; findings should therefore be interpreted within the context of each outcome domain.

To assess perturbation effects in the proximal task (NewMap vs NewSeq) at DROP, z-scored Speed, Accuracy and BCF were analyzed using LMM. Pairwise estimated marginal means (EMMs) quantified subgroup-specific DROP effects, and improvement over T31–T40 was analyzed using additional LMM.

For proximal–distal comparisons, participant-specific intercepts and slopes extracted from LMM were used as estimates of baseline performance and improvement for each metric. Group differences in improvement (T1-T30) were assessed using linear regression models adjusted for estimated baseline performance. To enable direct comparisons between proximal and distal measures, the z-scored proximal metrics (Speed, Accuracy, and BCF) were combined and compared with the combined z-scored distal metrics (Dext-Accuracy and BICO) using a linear model.

Multiple linear regression models examined associations between individual learning improvement in stroke patients and baseline demographics, cognitive and motor impairments. Individual learning slopes and estimated baseline performances were extracted from LMM including patient-specific random intercepts and random slopes for training improvement. Multicollinearity among predictors was assessed using variance inflation factors (VIF). Because FMA-UE, AMAT, and SAFE scores showed substantial multicollinearity^35^, they were combined into a composite motor score (Motor_PC1) using principal component analysis (PCA). Standardized regression coefficients (β) with 95% confidence intervals were used to quantify association effects.

## Results

### Participants characteristics

Between April 2023 and December 2025, 593 stroke patients were screened, 121 were eligible and 109 patients participated in the study (Fig. S2). Additionally, 62 age-matched HI were recruited. Patients and HI did not differ significantly in age (67.6±13.2 versus 64.7±8.0 years, respectively; p=0.13). Twelve patients were excluded because of deteriorating health, multiple lesions identified on MRI, or premature hospital discharge. Two HI were excluded following identification of a neurological disorder (early dementia and a previous stroke).

Overall, patients exhibited mild stroke severity (NIHSS=2.9±2.0), moderate UL motor impairment (FMA-UE=56.5±14.4) and mild sensory impairment (FMA-UE sensation=11.6±1.4), and mild cognitive deficits (MoCA=22.4±5.2). UL impairment was severe in 7 patients (FMA-UE <30), moderate in 9 (31-45), and mild in 81 (>45). Cognitive impairment was classified as moderate in 19 patients (MoCA :10-17), mild in 40 (18-25), and absent in 34 (26-30). Thirty HI and 45 patients were randomized to the NewSeq condition, whereas 30 HI and 52 patients were randomized to the NewMap condition (Table 1).

**Table 1.**
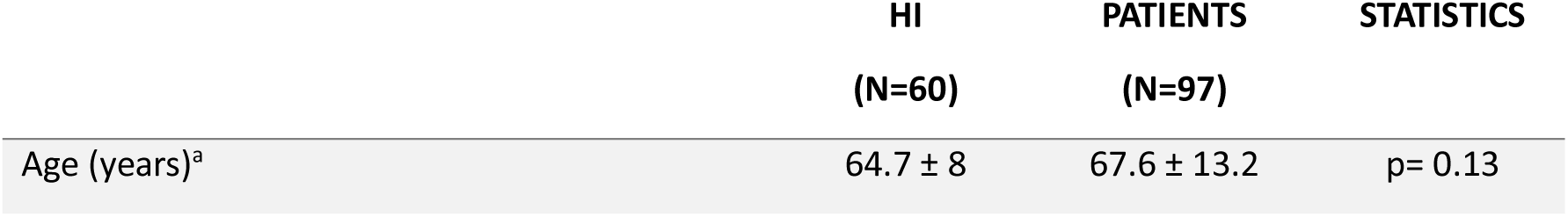
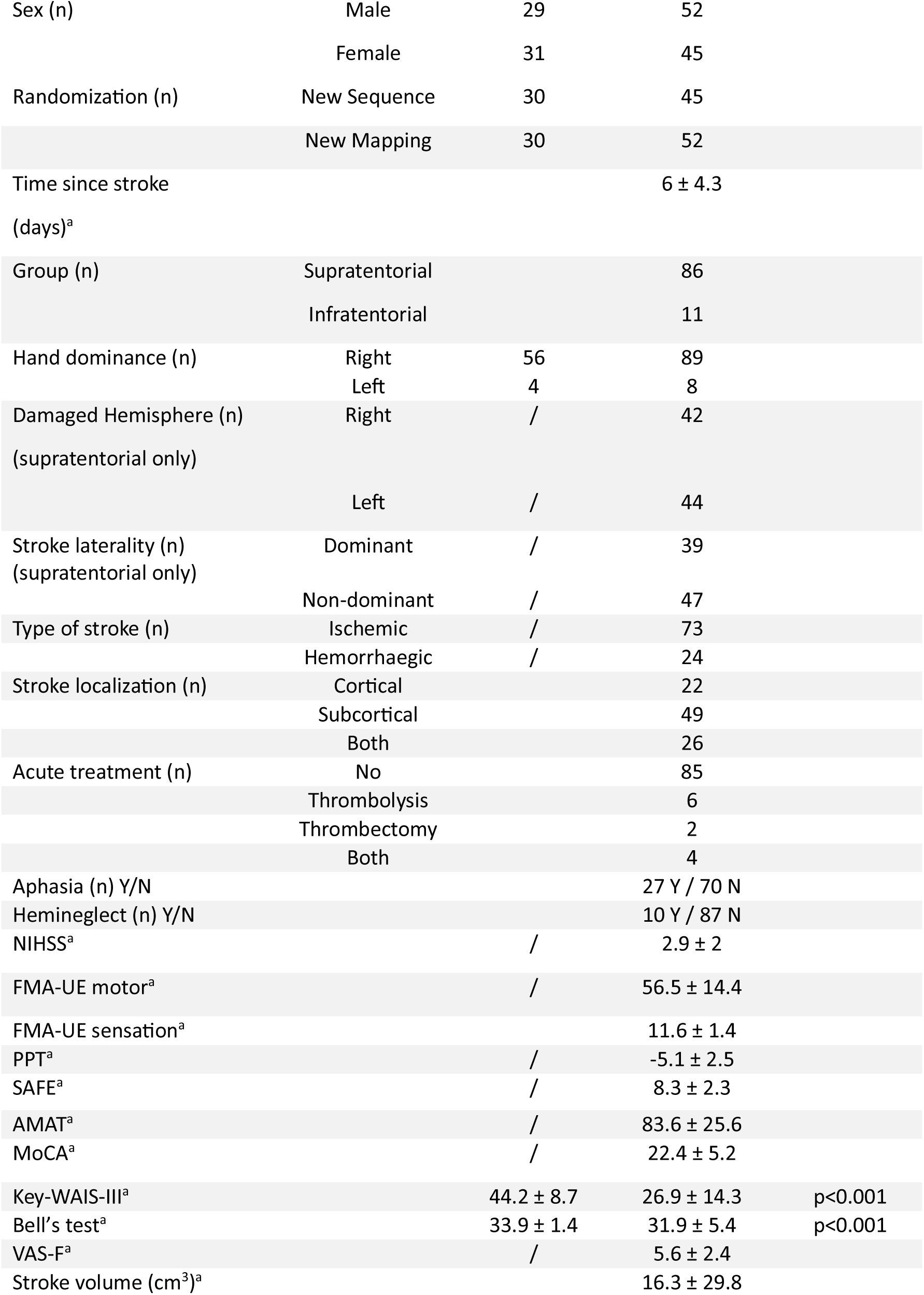
Demographics. NIHSS: NIH Stroke Scale, FMA-UE: Fugl-Meyer Assessment – Upper Extremity, PPT: Purdue Pegboard Test, SAFE: shoulder Abduction Finger Extension, AMAT: Arm Motor Ability Test, MoCA: Montreal Cognitive Assessment, Key-WAIS-III: Wechsler Adult Intelligence Scale-Key subtest, VAS-F: Visual Analogue Scale-Fatigue. ^a^ mean±SD.

|  | HI<br>(N=60) | PATIENTS<br>(N=97) | STATISTICS |
| --- | --- | --- | --- |
| Age (years)<sup>a</sup> | 64.7 ± 8 | 67.6 ± 13.2 | p= 0.13 |

**Table 1. Demographics.** NIHSS: NIH Stroke Scale, FMA-UE: Fugl-Meyer Assessment – Upper Extremity, PPT: Purdue Pegboard Test, SAFE: shoulder Abduction Finger Extension, AMAT:
|  |  |  |  |  |
| --- | --- | --- | --- | --- |
| Sex (n) | Male | 29 | 52 |  |
|  | Female | 31 | 45 |  |
| Randomization (n) | New Sequence | 30 | 45 |  |
|  | New Mapping | 30 | 52 |  |
| Time since stroke (days)<sup>a</sup> |  |  | 6 ± 4.3 |  |
| Group (n) | Supratentorial |  | 86 |  |
|  | Infratentorial |  | 11 |  |
| Hand dominance (n) | Right | 56 | 89 |  |
|  | Left | 4 | 8 |  |
| Damaged Hemisphere (n) (supratentorial only) | Right | / | 42 |  |
|  | Left | / | 44 |  |
| Stroke laterality (n) (supratentorial only) | Dominant | / | 39 |  |
|  | Non-dominant | / | 47 |  |
| Type of stroke (n) | Ischemic | / | 73 |  |
|  | Hemorrhagic | / | 24 |  |
| Stroke localization (n) | Cortical |  | 22 |  |
|  | Subcortical |  | 49 |  |
|  | Both |  | 26 |  |
| Acute treatment (n) | No |  | 85 |  |
|  | Thrombolysis |  | 6 |  |
|  | Thrombectomy |  | 2 |  |
|  | Both |  | 4 |  |
| Aphasia (n) Y/N |  |  | 27 Y / 70 N |  |
| Hemineglect (n) Y/N |  |  | 10 Y / 87 N |  |
| NIHSS<sup>a</sup> |  | / | 2.9 ± 2 |  |
| FMA-UE motor<sup>a</sup> |  | / | 56.5 ± 14.4 |  |
| FMA-UE sensation<sup>a</sup> |  |  | 11.6 ± 1.4 |  |
| PPT<sup>a</sup> |  | / | -5.1 ± 2.5 |  |
| SAFE<sup>a</sup> |  | / | 8.3 ± 2.3 |  |
| AMAT<sup>a</sup> |  | / | 83.6 ± 25.6 |  |
| MoCA<sup>a</sup> |  | / | 22.4 ± 5.2 |  |
| Key-WAIS-III<sup>a</sup> |  | 44.2 ± 8.7 | 26.9 ± 14.3 | p<0.001 |
| Bell's test<sup>a</sup> |  | 33.9 ± 1.4 | 31.9 ± 5.4 | p<0.001 |
| VAS-F<sup>a</sup> |  | / | 5.6 ± 2.4 |  |
| Stroke volume (cm<sup>3</sup>)<sup>a</sup> |  |  | 16.3 ± 29.8 |  |

### Proximal bim-MSkL

Unexpectedly, patients exhibited higher baseline Speed than HI (β=0.66, SE=0.20, p=0.001). However, Speed in patients decreased markedly during the first three training blocks (Fig. 2), and they remained significantly slower than HI at T30 (β=-1.42, SE=0.27, p<0.001). Across the three training days, patients demonstrated smaller Speed gains than HI (group × training block interaction: β=-0.038, SE=0.006, p<0.001).

**Figure 2.**
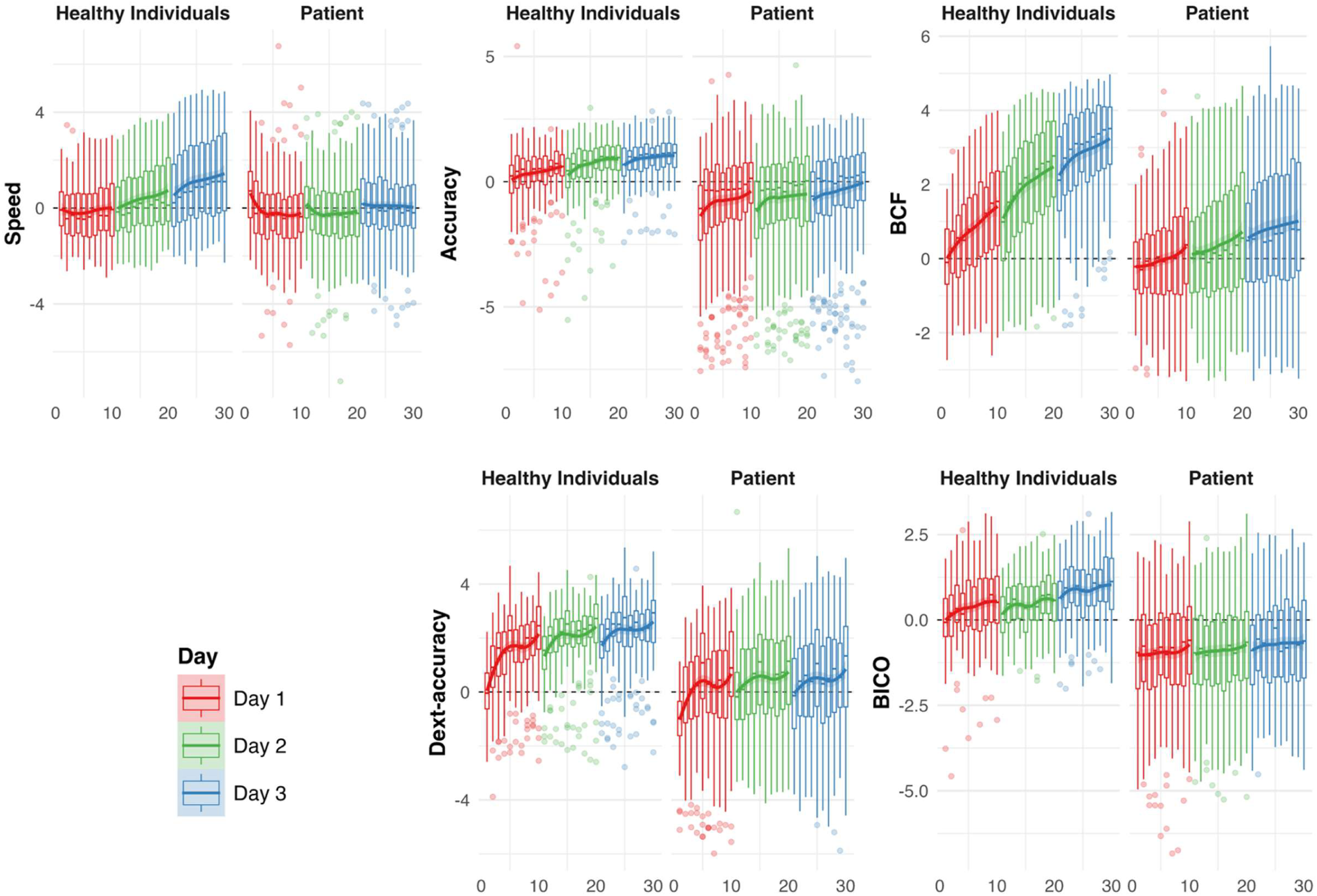
Z-score progression across training blocks. For each of the five metrics of interest of the proximal and distal tasks, the progression over D1-D2-D3 for HI and patients was plotted in boxplots. X axes represent training blocks (1-30). Y axes represent metrics of interest in z-scores with T1 in HI as the reference. The boxplots represent the median and interquartile range (IQR). Vertical whiskers extend to values within 1.5*IQR. Training blocks of D1 (T1-T10) in red, of D2 (T11-T20) in green and of D3 (T21-T30) in blue. Bold lines represent loess-smoothed trend lines with 95% CI. Data were visualized across training blocks using loess-smoothed trend lines with 95% confidence intervals (CI).

HI exhibited minimal overnight changes in Speed between training days (overnight retention) and continued to improve gradually across sessions (D1-D2: β=0.014, SE=0.13, p=0.92; D2-D3: β=0.24, SE=0.12, p=0.05). In contrast, patients began D2 and D3 at higher speeds than those achieved at the end of the previous day (D1-D2: β=-0.37, SE=0.17, p=0.03; D2-D3: β=-0.51, SE=0.16, p=0.002), followed by progressive slowing across subsequent training blocks.

When the first three blocks of D1 were excluded (T1-T3), baseline Speed no longer differed between groups (β=-0.14, SE=0.21, p=0.50), whereas the group difference in learning slope remained significant (β=-0.034, SE=0.007, p<0.001; Fig. S3).

Patients exhibited lower Accuracy than HI at both baseline (β=-1.40, SE=0.27, p<0.001) and T30 (β=-1.06, SE=0.24, p<0.001). However, Accuracy improvement did not differ significantly between groups (group × training block interaction: β=-0.004, SE=0.005, p=0.42). Patients showed a larger overnight decline in Accuracy between D1 and D2 than HI (HI: β=0.37, SE=0.17, p=0.04; patients: β=0.60, SE=0.23, p=0.01), whereas overnight changes between D2 and D3 were not significant in either group (HI: β=0.25, SE=0.15, p=0.09; patients: β=0.06, SE=0.19, p=0.70). Nevertheless, both groups retained learning gains across training days (Fig. 2).

For BCF, baseline performance did not differ significantly between patients and HI (β=-0.20, SE=0.16, p=0.21). However, patients exhibited lower coordination than HI at T30 (β=-2.20, SE=0.28, p<0.001). Although BCF improved in both groups, improvement was attenuated in patients (group × training block interaction: β=-0.040, SE=0.006, p<0.001). HI showed small but significant overnight declines in BCF (D1-D2: β=0.27, SE=0.12, p=0.03; D2-D3: β=0.25, SE=0.10, p=0.04), whereas overnight changes were not significant in patients (D1-D2: β=-0.03, SE=0.16, p=0.84; D2-D3: β=-0.14, SE=0.15, p=0.35).

### Distal bim-MSkL

For Dext-Accuracy, patients performed worse than HI at baseline (β=-1.05, SE=0.19, p<0.001) and at T30 (β=-1.74, SE=0.32, p<0.001). Both groups improved with practice, although learning was reduced in patients (group × training block interaction: β=-0.020, SE=0.007, p=0.02). HI exhibited significant overnight declines between training days (D1-D2: β=0.89, SE=0.25, p<0.001; D2-D3: β=0.71, SE=0.22, p=0.002), but performance remained above baseline levels. Overnight retention did not differ significantly between patients and HI (D1-D2: β=-0.16, SE=0.31, p=0.60; D2-D3: β=0.11, SE=0.28, p=0.68; Fig. 2).

Similarly, patients exhibited poorer BICO performance than HI at baseline (β=-1.03, SE=0.21, p<0.001) and T30 (β=-1.64, SE=0.23, p<0.001). Improvement was significantly smaller in patients than in HI (group × training block interaction: β=-0.010, SE=0.006, p=0.02). No significant overnight changes were observed for either HI (D1-D2: β=0.35, SE=0.18, p=0.055; D2-D3: β=-0.09, SE=0.17, p=0.57) or patients (D1-D2: β=-0.015, SE=0.23, p=0.94; D2-D3: β=0.32, SE=0.21, p=0.14).

### Control policy versus motor sequence learning

For Speed, neither the NewMap nor NewSeq perturbation induced a significant performance drop in HI (NewMap: β=0.04, SE=0.18, p=0.80; NewSeq: β=-0.04, SE=0.18, p=0.80). In contrast, Speed increased transiently at the perturbation point in patients assigned to both NewMap (β=0.78, SE=0.14, p<0.001) and NewSeq (β=0.53, SE=0.15, p<0.001; Fig. 3). The magnitude of the DROP did not differ between NewMap and NewSeq in either HI (contrast=-0.08, SE=0.25, p=0.74) or patients (contrast=-0.17, SE=0.32, p=0.60). Relearning slopes did not differ significantly between NewMap/NewSeq patient subgroups (β=-0.04, SE=0.03, p=0.23). A group difference in relearning was observed only for the NewSeq condition, with patients showing smaller gains than HI (β=-1.06, SE=0.46, p=0.02).

**Figure 3.**
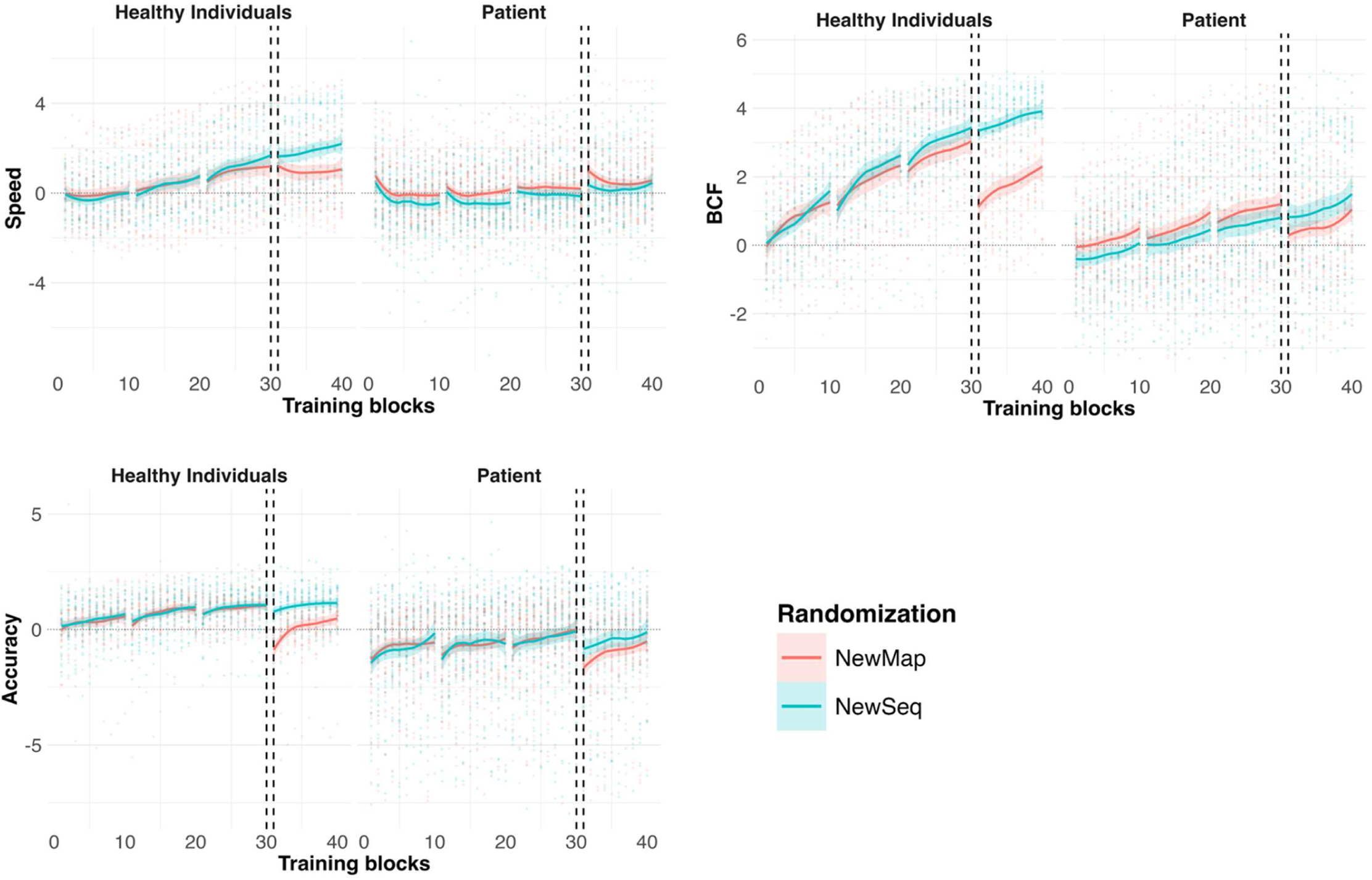
Effect of two generalization modes on the proximal bimanual task. Accuracy and Speed (upper row) and BCF (lower row) are plotted across groups (HI and patients) and randomization subgroups (NewMap in red and NewSeq in blue) over the three days. For each randomization subgroup, mean and 95% CI progression over the training blocks are plotted. X axes represent training blocks and Y-axes the metrics in z-score. After the 10 training blocks on D3 (vertical dotted lines), the randomization subgroups differed by their generalization modes for ten additional training blocks (T31-T40). In the NewMap group, the control policy was reversed by swapping which UL controlled the L-R displacement of the common cursor. In the NewSeq group, the *CIRCUIT* was rotated by 90°, changing the motor sequence while keeping the same control policy.

Accuracy dropped significantly following NewMap in both HI (β=1.89, SE=0.26, p<0.001) and patients (β=1.62, SE=0.21, p<0.001), whereas NewSeq induced a smaller decline that reached significance only in patients (β=0.66, SE=0.22, p=0.003) but not in HI (β=0.32, SE=0.26, p=0.22). The DROP was significantly larger in NewMap than NewSeq in both HI (contrast=1.57, SE=0.36, p<0.001) and patients (contrast=0.96, SE=0.30, p=0.002). In HI, relearning was slower in the NewSeq than the NewMap condition (β=-0.087, p=0.001). No such difference was observed in patients (β=0.021, SE=0.03, p=0.54). Relearning slopes did not differ significantly between HI and patients in either condition.

For BCF, NewMap induced a significant DROP in both HI (β=1.91, SE=0.17, p<0.001) and patients (β=0.93, SE=0.14, p<0.001), whereas NewSeq did not significantly affect performance (HI: β=0.11, SE=0.17, p=0.53; patients: β=-0.15, SE=0.14, p=0.29). The DROP was significantly larger for NewMap than NewSeq in both HI (contrast=1.80, SE=0.24, p<0.001) and patients (contrast=1.08, SE=0.20, p<0.001). Relearning was slower in the HI NewSeq subgroup than in the HI NewMap subgroup (β=-0.051, SE=0.03, p=0.044), whereas no subgroup difference was observed in patients (β=0.03, SE=0.03, p=0.35). Compared with HI, patients showed significantly reduced relearning only in the NewMap condition (β=-1.61, SE=0.44, p<0.001).

### Proximal versus distal task

HI demonstrated greater bim-MSkL on the proximal than on the distal task (β=0.03, p<0.001), consistent with a proximal-distal learning gradient. Patients showed better baseline performance on the proximal than on the distal task (β=0.74, p<0.001), but no significant difference in learning between tasks (β=-0.01, SE=0.008, p=0.23; Fig. 4).

**Figure 4.**
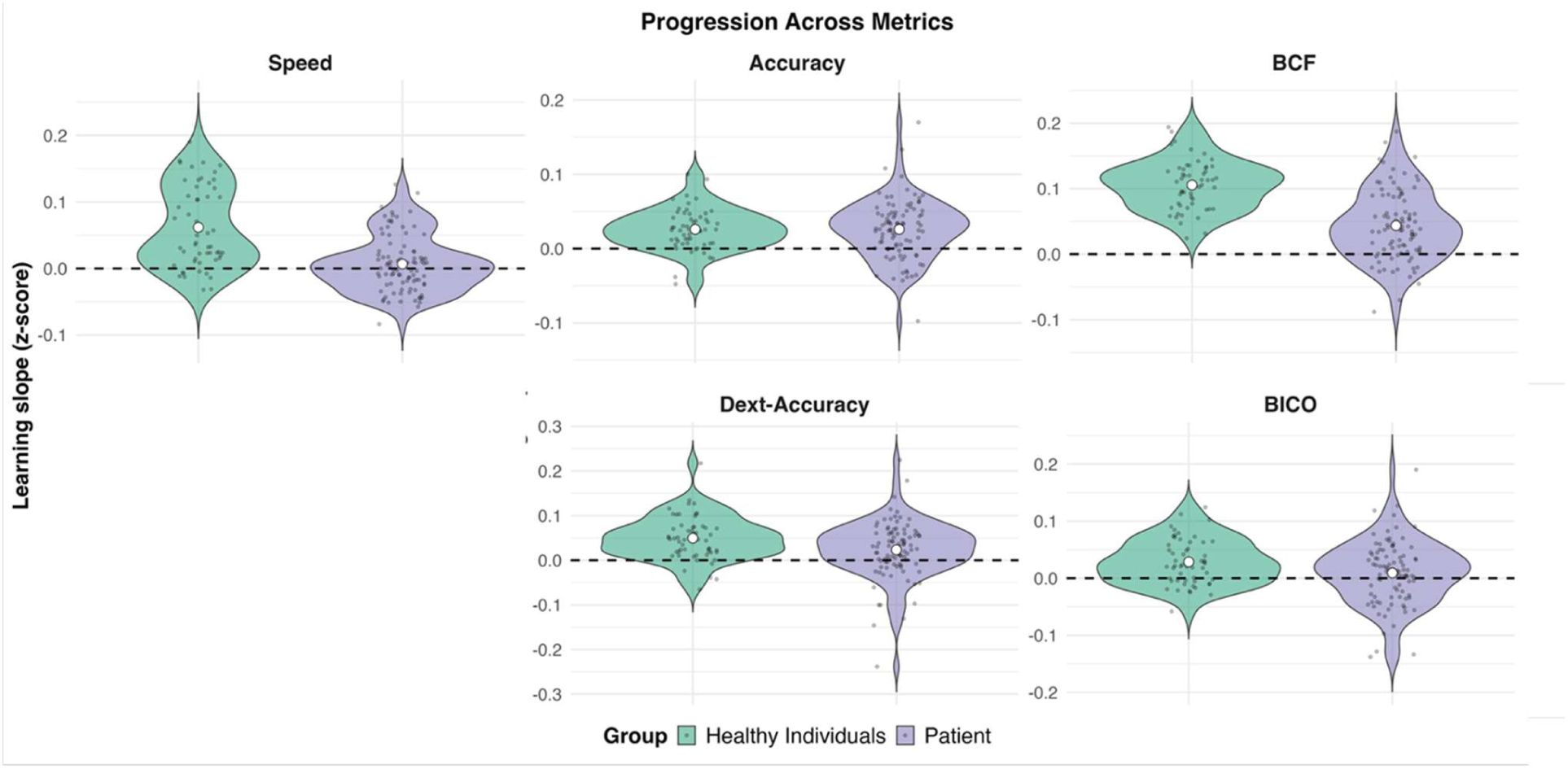
Comparison between proximal and distal task progression. For each metric, HI (green) and patient’s (violet) violin plots represent the distribution of participants progression (T1-T30) expressed in z-score. The horizontal dashed lines correspond to the HI’s means at baseline (T1), used as references for computing z-scores. Violins represent the kernel density of progression (learning slopes). The width of the violins represents the density of participants. Each dot represents one individual. The white point represents the group’s mean learning slope. Upper row: proximal task; lower row: distal task.

Compared with HI, patients exhibited poorer baseline performance on both the distal and proximal tasks ((β=-1.04, SE=0.15, p<0.001; Fig. S4)). Across both tasks, patients demonstrated smaller learning-related improvements than HI (β=-0.03, SE=0.006, p<0.001). This pattern was observed for all outcome measures (all p<0.001) except proximal Accuracy (β=-0.007, SE=0.007, p=0.26).

### Multiple regression models

For Speed, the regression model explained 17% of the variance early after stroke (adjusted R²=0.17, p<0.001). Key WAIS-III performance was positively associated with Speed improvement (β=0.42, SE=0.18, p=0.02; Fig. 5).

**Figure 5.**
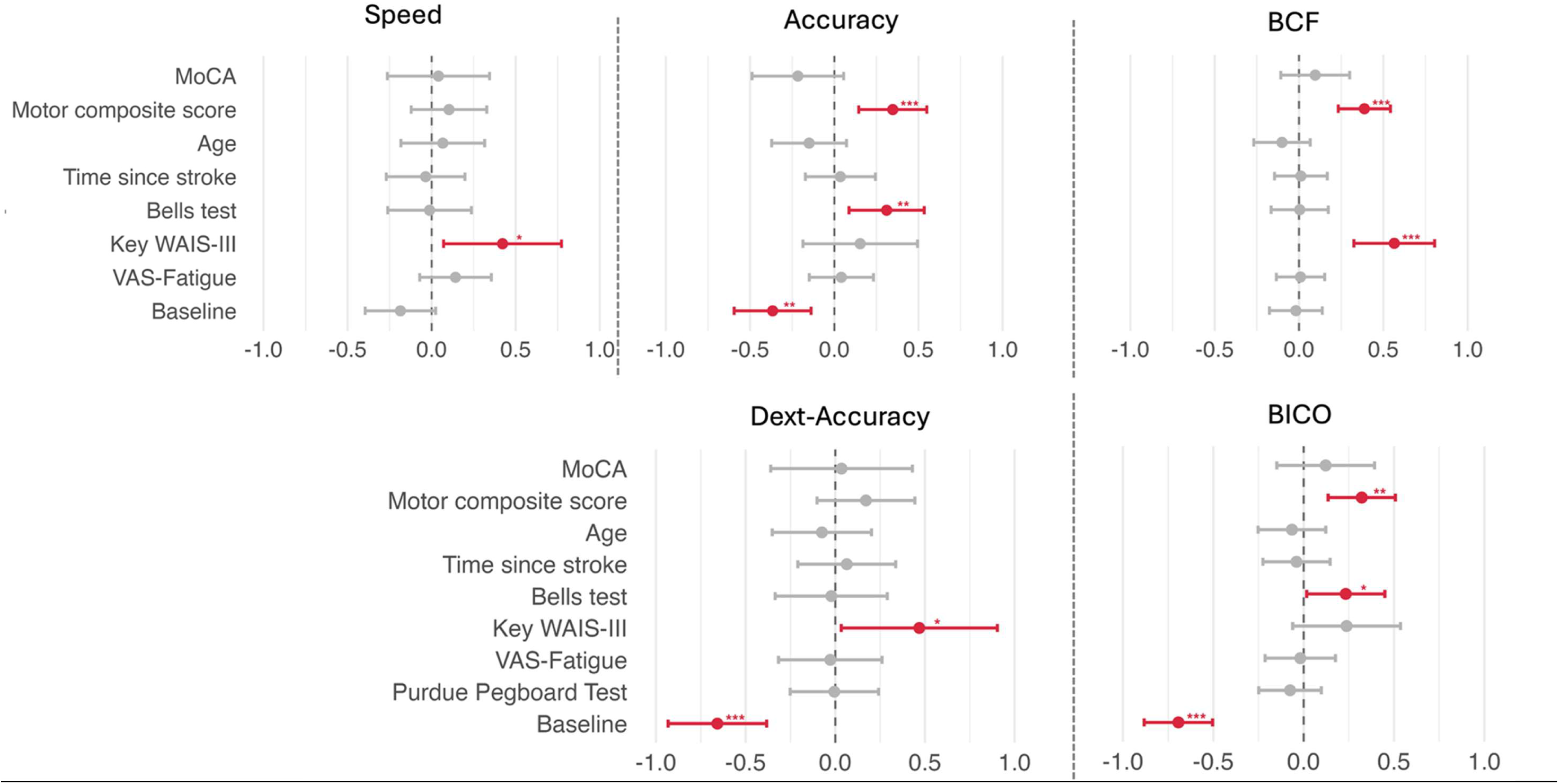
Multiple regression models in acute stroke patients. For each metric, multiple regression models are plotted using demographic (age) and clinical predictors. For each predictor, the standardized regression coefficient or β (point) and the 95% confidence interval (horizontal bar) are plotted. Significant predictors in red. * p<0.05, ** p<0.01, *** p<0.001. Upper row: proximal task; lower row: distal task.

For Accuracy, the model explained 22% of the variance (adjusted R²=0.22, p<0.001). Accuracy improvement was negatively associated with baseline Accuracy (β=-0.37, SE=0.11, p=0.002) and positively associated with Motor_PC1 (β=0.35, SE=0.10, p<0.001) and Bells Test performance (β=0.31, SE=0.11, p=0.006).

For BCF, the model explained 64% of the variance (adjusted R²=0.64, p<0.001). Both Motor_PC1 (β=0.39, SE=0.08, p<0.001) and Key WAIS-III performance (β=0.56, SE=0.12, p<0.001) were positively associated with improvement.

For Dext-Accuracy, the model explained 27% of the variance (adjusted R²=0.27, p=0.002). Improvement was positively associated with Key WAIS-III performance (β=0.47, SE=0.22, p=0.04) and negatively associated with baseline Dext-Accuracy (β=-0.66, SE=0.14, p<0.001).

For BICO, the model explained 54% of the variance (adjusted R²=0.54, p<0.001). BICO improvement was negatively associated with baseline BICO (β=-0.69, SE=0.09, p<0.001) and positively associated with both Motor_PC1 (β=0.32, SE=0.09, p=0.001) and Bells Test performance (β=0.23, SE=0.11, p=0.04).

Residual diagnostics revealed no major deviations from normality and no evidence of substantial nonlinear relationships.

## Discussion

Patients with mild-to-moderate (sub)acute stroke retained substantial, albeit attenuated, capacity to acquire novel proximal and distal bimanual motor skills relative to HI. In both groups, altering the control policy produced larger performance disruptions than changing the motor sequence, highlighting the prominent contribution of sensorimotor mappings to early bimanual skill acquisition. In patients, however, adaptation to changes in the established control policy appeared less flexible. Furthermore, motor and attentional capacities were significant predictors of learning-related improvements, indicating that residual motor and cognitive resources influence bim-MSkL after stroke.

### Preserved bim-MSkL in subacute stroke

Patients exhibited impaired performance on both proximal and distal tasks at baseline, reflecting impairments in motor control. Nevertheless, they demonstrated clear learning-related improvements and overnight retention across training, indicating that acquisition of novel coordinated sensorimotor skills remains possible during the subacute phase after stroke^36,37^. Consistent with previous studies, HI showed progressive gains in speed, accuracy, and coordination over three days of training^8,20,30,31^. Although learning-related improvements were attenuated in patients compared with HI, the overall pattern supports the persistence of substantial bimanual motor learning capacity despite neurological impairment as demonstrated beforehand in chronic stroke patients^8,30,31^.

Interestingly, patients initially performed the proximal task at higher speeds than HI but with markedly reduced accuracy. During the first training blocks, speed declined rapidly while accuracy improved substantially. This pattern could likely reflect rapid strategic recalibration to the robotic environment and task demands. Subsequently, proximal accuracy continued to improve steadily, indicating retained error-reduction capacity despite impaired motor execution^38^.

The greater gains in accuracy than in speed in patients suggest a shift in the speed-accuracy trade-off, potentially reflecting either compensatory strategies or stroke-related limitations in force generation and movement velocity^39^. Coordination improved more gradually but consistently, despite impaired baseline interlimb control^40^. Notably, coordination and accuracy improvements were only partially coupled in patients, suggesting that interlimb coordination and error reduction may rely on at least partially distinct learning processes^30,31^.

Similar findings were observed for distal bimanual motor learning. Despite impaired baseline dexterous performance, patients improved both dexterous accuracy and bimanual coordination over training, although to a lesser extent than HI. The preservation of overnight retention further supports the ability of subacute stroke patients to acquire and retain novel bimanual dexterous sensorimotor skills.

### Control policy and motor sequence

In HI, reversing the learned bimanual control policy produced larger performance disruptions than changing the motor sequence, consistent with previous evidence suggesting that early bimanual motor learning is driven primarily by acquisition of a control policy, which would dominate over sequence-specific refinements^20^.

A comparable pattern was observed in patients for accuracy and coordination outcomes, with larger performance decrements following control-policy perturbation than sequence changes. In contrast, speed increased transiently immediately after perturbation and then quickly declined, suggesting the use of a compensatory strategy. In HI, the larger disruption associated with control-policy changes was accompanied by steeper relearning slopes. This distinction was absent in patients, whose relearning rates did not differ between perturbation conditions. One interpretation is that stroke reduces the flexibility of internal sensorimotor representations, limiting selective updating of previously learned control policies. Alternatively, patients may rely on compensatory coordination strategies that generalize across perturbations, thereby diminishing behavioral differences between conditions^41^.

### No clear proximal-distal gradient in subacute stroke

HI exhibited greater learning-related improvements on the proximal task than on the distal task, indicating a proximal-distal gradient in bim-MSkL with these two tasks. In contrast, patients showed comparable learning rates across proximal and distal tasks despite markedly poorer distal baseline performance. Thus, although distal motor control was more severely impaired, distal deficits did not translate into disproportionately impaired learning capacity. The absence of a proximal-distal learning gradient early after stroke is noteworthy because proximal and distal UL segments rely on partially distinct neural substrates. Although no consensus currently exists regarding proximal-distal patterns of post-stroke recovery^16,42^, our findings suggest that learning of complex bimanual skills may be constrained by shared network-level mechanisms rather than by segment-specific impairments alone^16^. Acute stroke-related impairments may therefore obscure the proximal-distal gradient observed in HI, leading both proximal and distal control processes to depend on common compensatory mechanisms. Heterogeneity in lesion location may also contribute to this effect. Lesions involving motor, premotor, or deep hemispheric structures may disproportionately affect proximal control, whereas lesions involving the primary motor cortex may produce relatively greater distal impairment^14^. Finally, differences in task structure may have influenced learning patterns, as the proximal task required asymmetric interlimb coordination whereas the distal task relied on symmetrical pinching forces and thus lower coordination demands^40^.

### Contributors of proximal and distal bim-MSkL in subacute stroke

Regression analyses identified motor and attentional impairments as important correlates of bimanual motor learning in subacute stroke (17-64% of variance), as suggested previously ^43,44^. In the proximal task, better motor function was associated with greater improvements in accuracy and coordination, suggesting that preserved motor control facilitates both acquisition and expression of learned motor behaviours^45,46^.

Likewise, better attentional and visuospatial capacities were associated with greater improvements in speed, accuracy and coordination, indicating that successful bimanual learning depends not only on motor execution but also on cognitive processes such as attentional engagement, working memory, and visuospatial monitoring^47^. The negative association between baseline accuracy and subsequent improvement likely reflects ceiling effects in participants with better initial performance.

In the distal task, attention and visuospatial planning were positively associated with accuracy and coordination improvement. Retained motor function was positively related to coordination gains, consistent with previous findings^48,49^. Baseline performances were negatively associated with both distal accuracy and coordination improvement, reflecting ceiling effects in both distal metrics.

Collectively, the convergence of predictors across proximal and distal tasks suggests that bimanual motor learning in different UL segments relies on shared functional resources involving both motor control and attentional processing. These findings support the view that motor and cognitive capacities jointly shape learning potential during the acute-subacute phase after stroke.

## Limitations

Several limitations should be acknowledged. First, the cohort predominantly comprised patients with mild-to-moderate stroke severity, with relatively few participants showing severe motor impairment, limiting generalizability to more severely affected populations. Second, the proximal and distal tasks differed substantially in their motor, cognitive, and coordination requirements, which may have reduced the sensitivity of direct proximal-distal comparisons despite z-score normalization. Third, although motor and attentional variables emerged as significant predictors of learning-related improvements, the proportion of explained variance remained moderate, suggesting that important determinants of bimanual motor learning early after stroke were not captured in the present analyses. Additional factors, including proprioceptive integrity, motivation, lesion location, and network-level brain characteristics, may further explain inter-individual variability^50^.

## Conclusion

Patients with mild-to-moderate subacute stroke retained substantial capacity for proximal and distal bimanual motor skill learning despite impaired baseline performance. Learning-related improvements were broadly comparable across proximal and distal UL segments and were associated with residual motor and attentional capacities. These findings support the potential value of early assessment of both motor and cognitive reserves when designing rehabilitation programs. Tailoring motor-learning interventions to individual patient capacities may enhance transfer to activities of daily living and optimize recovery during the early post-stroke period of heightened neuroplasticity.

## Data Availability

The data will we available upon request.

## Acknowledgements

We thank the patients and healthy individuals who kindly participated in this study. We thank the Stroke Unit team and Physical Medicine & Rehabilitation Department of the CHU UCL Namur - Godinne site for their implication, expertise and collaboration. We thank Maxime Teremetz and Steven Puisas from the Dextrain company (France) for co-developing the LIFT THE TRAY task.

## Abbreviations

MSkL: Motor skill learning
ADLs: Activities of daily living
Bim-MSkL: Bimanual motor skill learning
UL: Upper limb
HI: Healthy individuals
MoCA: Montreal cognitive assessment
NIHSS: National Institute of Health Stroke Scale
FMA-UE: Fugl-Meyer Assessment-Upper Extremity
SAFE: Shoulder Abduction Finger Extension
AMAT: Arm Motor Ability Test
PPT: Purdue Pegboard Test
WAIS: Wechsler Adult Intelligence Scale
ROM: Range of motion
NewMap: New Mapping
NewSeq: New Sequence
BCF: Bimanual coordination factor (proximal)
BICO: Bimanual coordination (distal)
Motor_PC1: Composite motor score
D1: Day 1
T1: Training block 1

